# Analysis of facial expressions recorded from patients during psychiatric interviews

**DOI:** 10.1101/2024.09.19.24313994

**Authors:** L. Mineur, M. Heide, S. Eickhoff, M. Avram, L. Franzen, F. Buschmann, F. Schröpfer, H.V. Rogg, C. Andreou, N.S. Brügge, H. Handels, S. Borgwardt, A. Korda

## Abstract

Mental health research increasingly focuses on the relationship between psychiatric symptoms and observable manifestations of the face and body^1^. In recent studies^2,3^, psychiatric patients have shown distinct patterns in movement, posture and facial expressions, suggesting these elements could enhance clinical diagnostics.

The analysis of the facial expressions is grounded on the Facial Action Coding System (FACS)^4^. FACS provides a systematic method for categorizing facial expressions based on specific muscle movements, enabling detailed analysis of emotional and communicative behaviors. This method combined with recent advancements in Artificial Intelligence (AI) has shown promising results for the detection of the patient mental state.

We analyze video data from patients with various psychiatric symptoms, using open-source Python toolboxes for facial expression and body movement analysis. These toolboxes facilitate face detection, facial landmark detection, emotion detection and motion recognition. Specifically, we aim to explore the connection between these physical expressions and established diagnostic tools, like symptom severity scores, and finally enhance psychiatric diagnostics by integrating AI-driven analysis of video data.

By providing a more objective and detailed understanding of psychiatric symptoms, this study could lead to earlier detection and more personalized treatment approaches, ultimately improving patient outcomes. The findings will contribute to the development of innovative diagnostic tools that are both efficient and accurate, addressing a critical need in mental health care.

## 1. Introduction

A crucial aspect of risk states in mental health is impairment in social function. Although the aetiopathology of this social function impairment is likely multifaceted, deficits in social communication may account for much of it. Mental health research is increasingly focusing on the relationship between psychiatric symptoms and observable manifestations of the face and body^1^. As a result, one must consider gestures and facial expression of emotion in the context of interpersonal interactions. Recent studies have shown that psychiatric patients exhibit distinct patterns of movement, posture and facial expression, suggesting that these elements could improve clinical diagnostics^2,3^. Automated measurement of face and head motion has provided the first objective measure of depression severity^5^. Video data have been applied in previous studies, which showed that significant differences in the facial expressions of emotion between clinical high-risk for psychosis patients and healthy controls can be identified in small fragments of behaviour; almost 1 minute^6,7^. Video analysis for the outcome prediction in psychiatry is still at a very early stage.

The analysis of facial expressions is based on the Facial Action Coding System (FACS). FACS provides a systematic method for categorizing facial expressions into Action Units (AUs). Each AU represents a distinct movement or combination of micro-expressions and voluntary movements, allowing researchers to describe and measure emotional states, non-verbal communication and facial behavior in a detailed and systematic way^4^.

Micro-expressions are fleeting and involuntary facial expressions that often occur in high-stakes situations when people try to cover up or hide their true emotions. These actions are controlled by the amygdala and are too brief (1/25 to 1/2 s) and inconspicuous for the natural eye to see^8^. In addition to the amygdala, other parts of the brain play an important role in the expression of emotions: Five cortical regions in each hemisphere are involved: primary motor cortex, ventral lateral premotor cortex, supplementary motor area, and rostral and caudal cingulate cortex^9^. The upper facial muscles receive bilateral cortical input, while the lower facial muscles predominantly receive contralateral input^10^.

The aim of our analysis is to try to predict depressive symptoms based on facial expressions. This method of using FACS, combined with recent advances in artificial intelligence (AI), has shown promising results in detecting the mental state of the patient^11^. We specifically try to find out the correlation between facial expressions and symptom severity, which are currently measured by questionnaires like the Beck’s Depression Inventory (BDI) score.

By providing a more objective and detailed understanding of depressive symptoms, the findings of this project could lead to earlier detection and more personalized treatment approaches, ultimately improving patient outcomes in individuals with depression. Given the high global prevalence and significant impact of depression on quality of life, there is a critical need for innovative diagnostic tools that are both efficient and accurate. The ability to predict and monitor depressive symptoms using objective data, such as facial expressions and speech patterns, could transform mental health care by enabling timely interventions and tailored treatments, offering a more reliable approach to managing depression.

## 2. Dataset and Method

### 2.1 Data

Our current dataset includes 58 videos from 32 different patients whose interviews with the clinician were recorded. Some of the patients come back for a re-evaluation or a follow-up appointment, but we used the videos as separate units. 22 of the videos are of female patients and 36 are of male patients. The average age of the patients is 32.71 years. 25 of the patients in the videos are taking some form of psychiatric medication, while 33 are either not taking any medication or are only taking non-psychiatric medication. The average BDI score is 18.40 (**Table 1**). The semi-structured interviews focused on depressive symptoms and are part of a structured schedule assessment in our hospital. The patients had received and filled out the self-assessment BDI before the interview with the clinician, which is a depression questionnaire with a scale between 0 and 63 points in order to assess the symptom severity^12^. 25 of the patients in the videos (43,1%) had already been diagnosed with an F33.X diagnosis, which according to ICD-10 is a recurrent depressive disorder. Thus, a balanced sample is used in this study.

**Table 1:** The mean values/distribution and standard deviation of the demographic and clinical variables of the analyzed dataset.

| #patients (videos) | age | sex | psychiatric medication | BDI score |
| --- | --- | --- | --- | --- |
| 32 (58) | 29.78 years<br>± 12,30 | 18 female<br>14 male | 13 yes<br>19 no | 18.40 ± 13,49 |

**Table 2:** Connection between AUs and Emotions based on *py-feat*.

| emotion | Associated AUs |
| --- | --- |
| anger | 4 + 5 + 7 + 23 |
| contempt | 6 + 12 + 14 |
| disgust | 6 + 9 + 11 + 15 + 17 |
| fear | 1 + 2 + 4 + 5 + 7 + 11 + 20 + 25 + 26 |
| happiness | 6 + 12 + 25 |
| sadness | 1 + 4 + 15 |
| surprise | 1 + 2 + 5 + 25 + 26 |

The videos were recorded using the Grasshopper3 USB3 camera with a lens that can record the micro-facial expressions bought from the Teledyne FLIR company.

The ethics committee of the University of Lübeck approved of the usage of this data for our project the 18^th^ July 2024 (No: 2024-355_1), and written informed consent is obtained from each participant. The study was conducted in accordance with the Declaration of Helsinki.

### 2.2 Pre-processing

First, we preprocess the videos by trimming them down to the parts where the patients speak about their depressive symptoms, which is the first part of the SCID interviews. Usually, the videos are about 5 to 15 minutes long.

For the actual analysis of the facial expressions, we use open-source python toolboxes like *py-feat*^13^. This is a freely available pre-trained model that can analyze video data frame by frame.

The components of *py-feat* include face detection, facial landmark detection and alignment, face and head pose estimation, action unit detection (AUs) and emotion detection. We focus on the action unit detection, as we try to find a correlation between the AUs and the BDI score. The different AUs are visualized in Figure 1 as they would look on a real face^14^.

**Figure 1:**
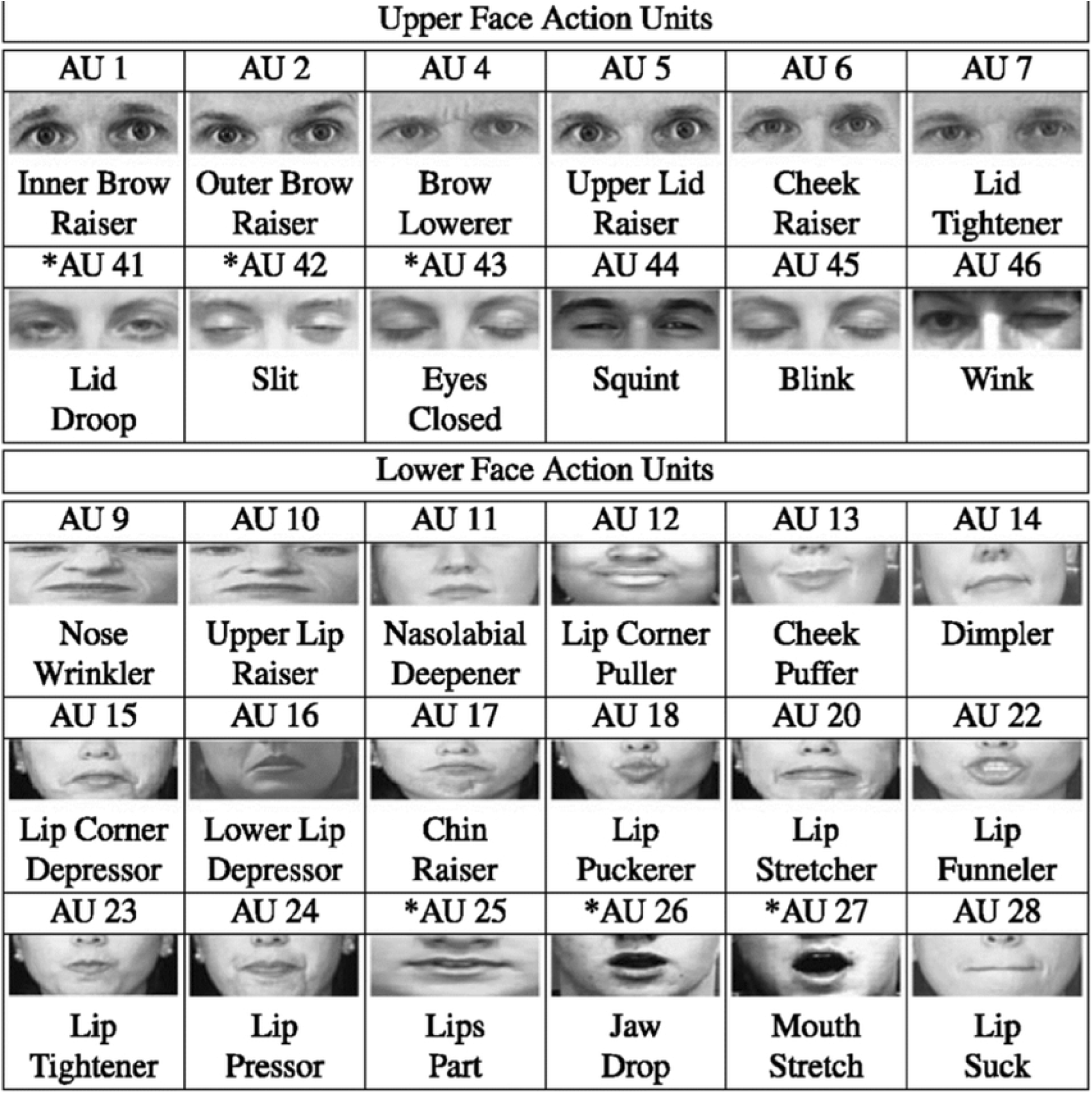
Upper and lower face AUs.

**Figure 2:**
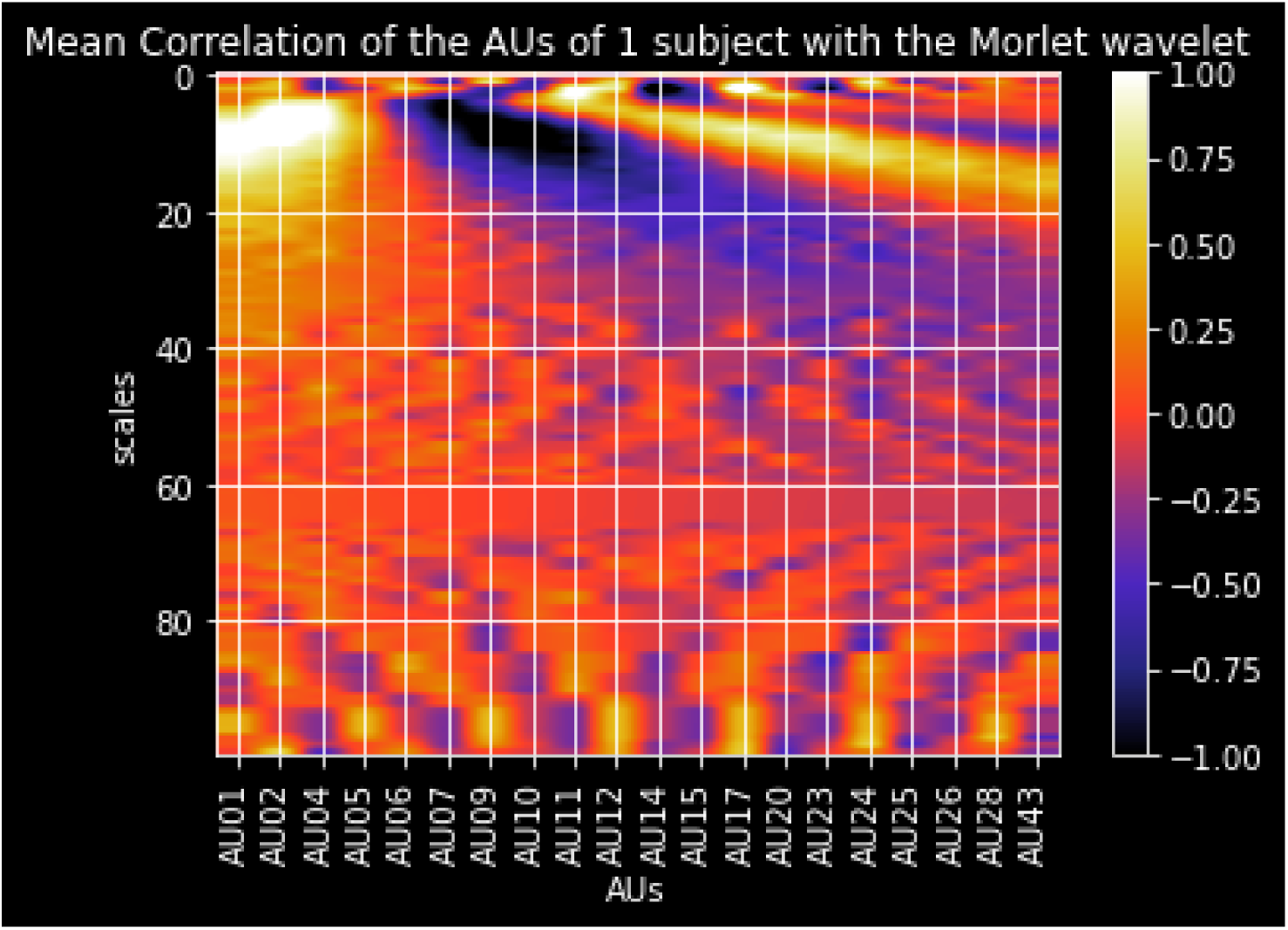
A 2D representation of the Aus.

### 2.3 Statistical analysis

We run a partial correlation analysis to estimate the association between the variables AUs and BDI score, with age, sex, type of camera, medication and a dummy variable for the videos coming from the same person as covariates of no interest. We calculate the FDR-corrected p-values to predict the BDI scores from the AUs.

### 2.4 Morlet Wavelet Transformation

The pipeline of the continuous wavelet transformation (CWT) followed a previous report by our group^15,16^. In short, CWT utilizes a fully scalable modulated window, offering a robust solution to the windowing function selection challenge in frequency-related (scale-related) signal processing methodologies^15^. In the present investigation, CWT was applied to decompose the AU series into frequency components for feature extraction. The CWT utilized the complex Morlet wavelet as its fundamental function to produce a spatial-scale representation of the original AU series, presenting it in the form of a “scalogram” plane. Within this plane, each individual value (referred to as a wavelet coefficient) signifies the degree of correlation between the AU series and the Morlet wavelet at specific point-scale pairs. The different scales (inverse frequencies) of the wavelet ranging from 0 to 100 were examined to identify the highest number of AUs with significant group differences.

## 3. Results

The python toolbox gives an output on which facial expressions and which emotions are expressed the most in the analyzed faces. Specifically, it gives us the average probability for the occurrence of each AU and emotion in the video we analyzed, while we only focused mainly on the AUs. An exemplary output is pictured by Cheong et al in Figure 3^13^.

**Figure 3:**
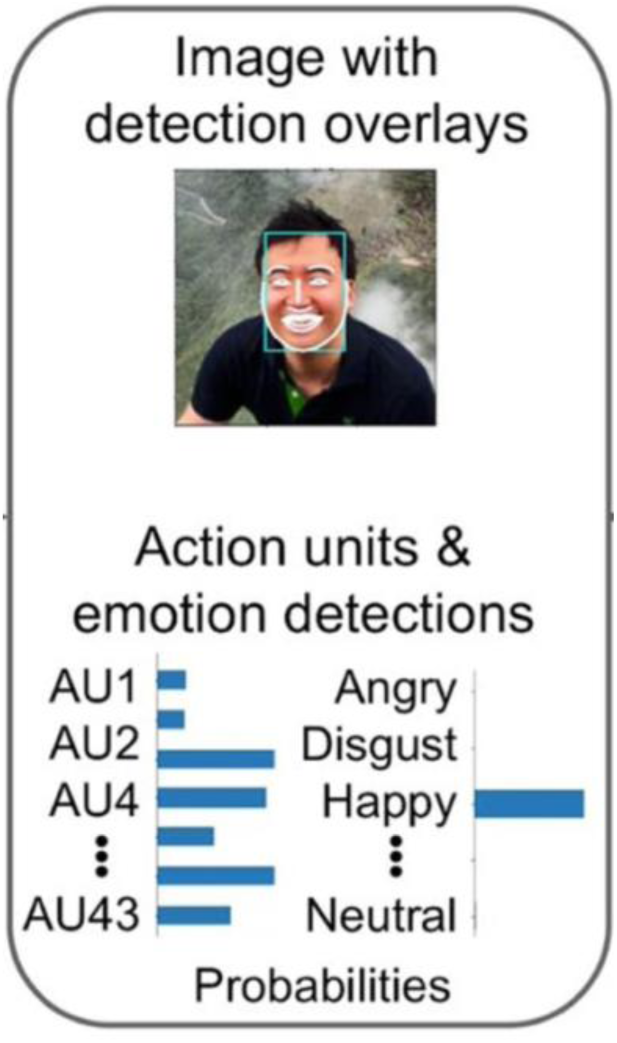
p*y-feat* output.

The partial correlation analysis showed a statistical significance, i. e. an FDR-corrected p-value below 0.05, for the **AUs 9, 11, 24, 25, 26** and **43**. Three of the AUs (9, 24 and 43) have a positive correlation with the BDI score, while the others (11, 25 and 26) have a negative coefficient, so inverse correlation with the BDI score. Results are presented in Table 3.

**Table 3:** partial correlation analysis results.

| AU | AU1 | AU2 | AU4 | AU5 | AU6 | AU7 | AU9 | AU10 | AU11 |
| --- | --- | --- | --- | --- | --- | --- | --- | --- | --- |
| **Coefficients** | -0.181 | 0.145 | 0.123 | 0.134 | 0.131 | 0.154 | 0.331* | -0.184 | -0.411* |
|  | **AU12** | **AU14** | **AU15** | **AU17** | **AU20** | **AU23** | **AU24** | **AU25** | **AU26** |
|  | -0.112 | -0.028 | -0.020 | 0.194 | -0.076 | -0.037 | 0.240* | -0.444* | -0.273* |
|  | **AU28** | **AU43** |  |  |  |  |  |  |  |
|  | -0.091 | 0.257* |  |  |  |  |  |  |  |

\*FDR corrected p-value $\leq 0.05$

These are the various AUs identified as particularly important in our analysis, including the facial muscles and emotions that are associated with them:

### AU9 – Nose Wrinkler

The muscle associated with AU9 is the levator labii superioris alaeque nasi muscle (Figure 4). It is a superficial muscle that lifts the upper lip and the nostrils. As specified by FACS, it is associated mainly with the emotion disgust. It appears to come up more the higher the BDI score is.

**Figure 4:**
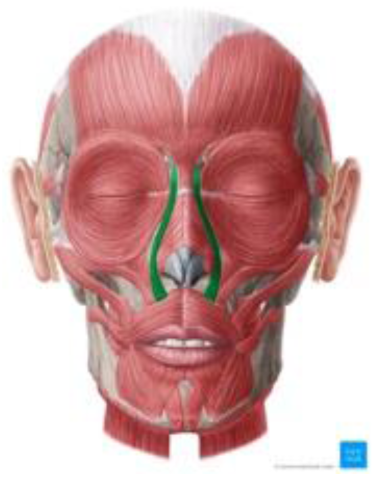
AU9 Nose Wrinkler. Image source: https://www.kenhub.com/de/library/anatomie/musculus-levator-labii-superioris-alaeque-nasi

### AU11 – Nasolabial Deepener

The AU 11 is characterized by the zygomaticus minor muscle (Figure 5). This muscle is a highly gracile, superficially located bundle of fibers^17^. It is originating from the lateral zygomatic arch to the skin of the medial upper lip and the orbicularis oris muscle. When contracted, the muscle elevates the upper lip, thus deepening the nasolabial furrow. FACS associates it with disgust and sadness and it is also called “the muscle of weeping and whimpering”^18^. In some studies with children and fetuses the deepening of the nasolabial furrow was associated with the expression of pain^19,20^. This AU is negatively associated with the depressive symptoms.

**Figure 5:**
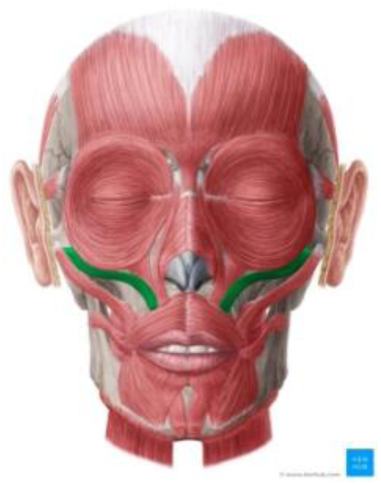
AU 11 Nasolabial Deepener. Image source: https://www.kenhub.com/de/library/anatomie/musculus-zygomaticus-minor

### AU24 – Lip Pressor

AU 24 is called the lip pressor and is characterized by the orbicularis oris muscle (Figure 6). This muscle is a complex, multi-layered muscle that is attached around the mouth and serves as an attachment site for other facial muscles^21^. As the name Lip pressor suggests, this AU represents the upper and lower lips being pressed against each other, thus the mouth being closed. This AU has a positive correlation with the BDI score.

**Figure 6:**
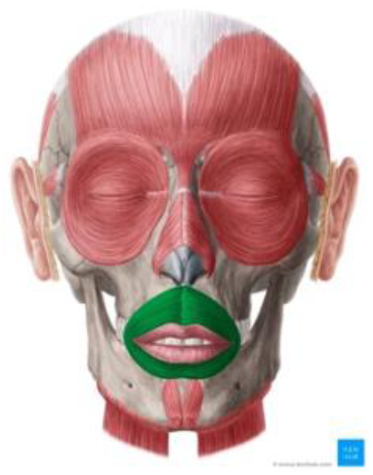
AU24 Lip Pressor. Image source: https://www.kenhub.com/de/library/anatomie/musculus-orbicularis-oris

### AU25 – Lip Part

This AU’s muscle is the depressor labii inferior muscle (Figure 7). It is a paired muscle that runs from the chin to the lower lip and, when contracted, pulls the lower lip down and forward. FACS associates this AU with happiness, surprise and fear. This seems to be expressed less the higher the BDI score is.

**Figure 7:**
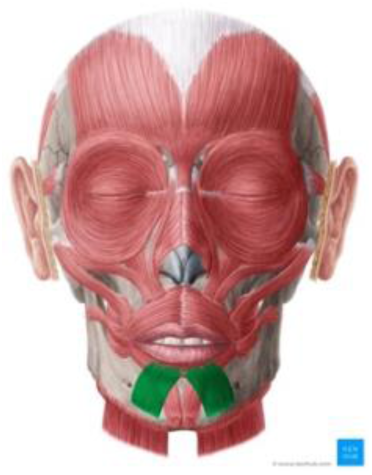
AU25 Lip Part. Image source: https://www.kenhub.com/de/library/anatomie/musculus-depressor-labii-inferioris

### AU26 – Jaw drop

This AU combines the masseter muscle, the temporal muscle and the medial pterygoid muscle (Figure 8). Through a sudden relaxation, these muscles produce the jaw drop, i.e. opening of the mouth. According to FACS, these muscles contribute to the emotions **fear and surprise**. This AU seems to be expressed less the higher the BDI score.

**Figure 8:**
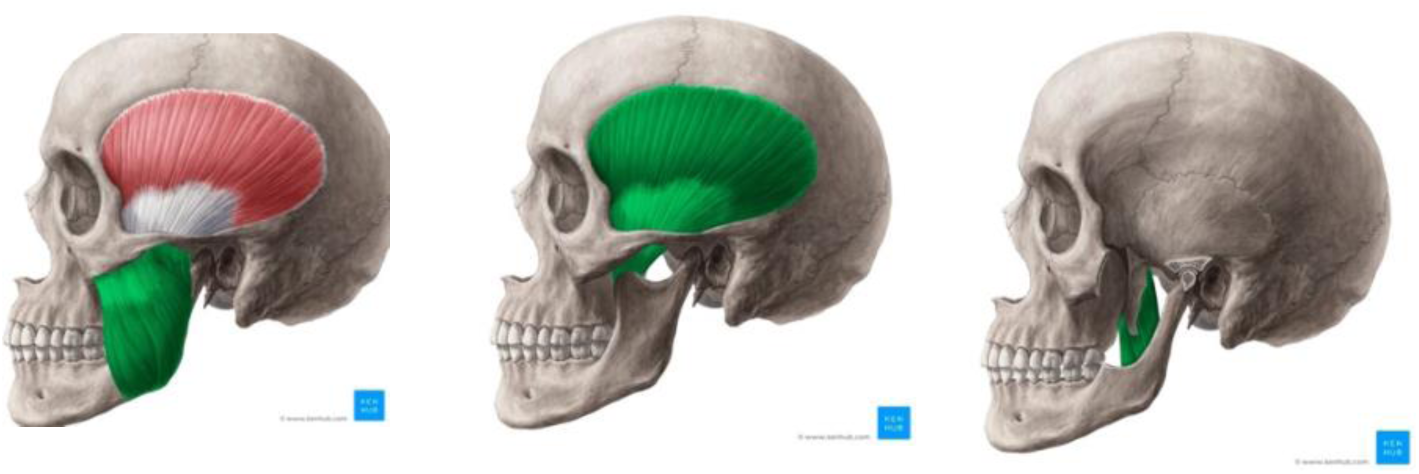
AU26 Jaw Drop. Image sources: https://www.kenhub.com/de/library/anatomie/musculus-masseter https://www.kenhub.com/de/library/anatomie/musculus-temporalis https://www.kenhub.com/en/library/anatomy/medial-pterygoid-muscle

### AU43 – Eyes Closed

The relevant muscle for the AU43 is the levator palpebrae superior muscle (Figure 9). In this case, the designated movement is not executed by contraction but relaxation of the muscle. This is one of the AUs from FACS that is **behavioral**, meaning that the person can easily target it intentionally, while the other mentioned AUs mostly happen unintentionally as an expression of emotion. This AU is positively associated with the BDI score.

**Figure 9:**
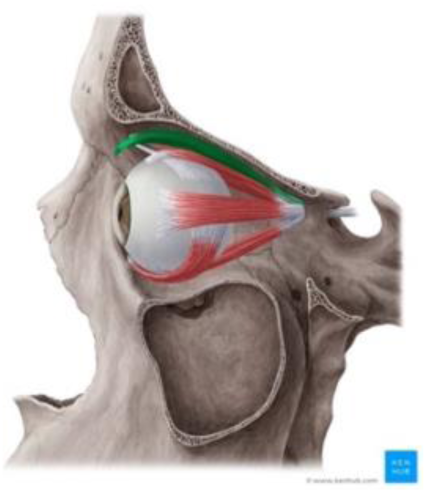
AU43 Eyes Closed. Image source: https://www.kenhub.com/de/library/anatomie/musculus-levator-palpebrae-superioris

The partial correlation analysis using the 2D representation of the AUs, as it is in Figure 2, showed a statistical significance, i. e. an FDR-corrected p-value below 0.05, for the **AUs 1, 6, 7, 9, 10, 11, 12, 14, 15, 17, 20, 23 and 25**. The correlation coefficients vary between negative and positive values for different scales.

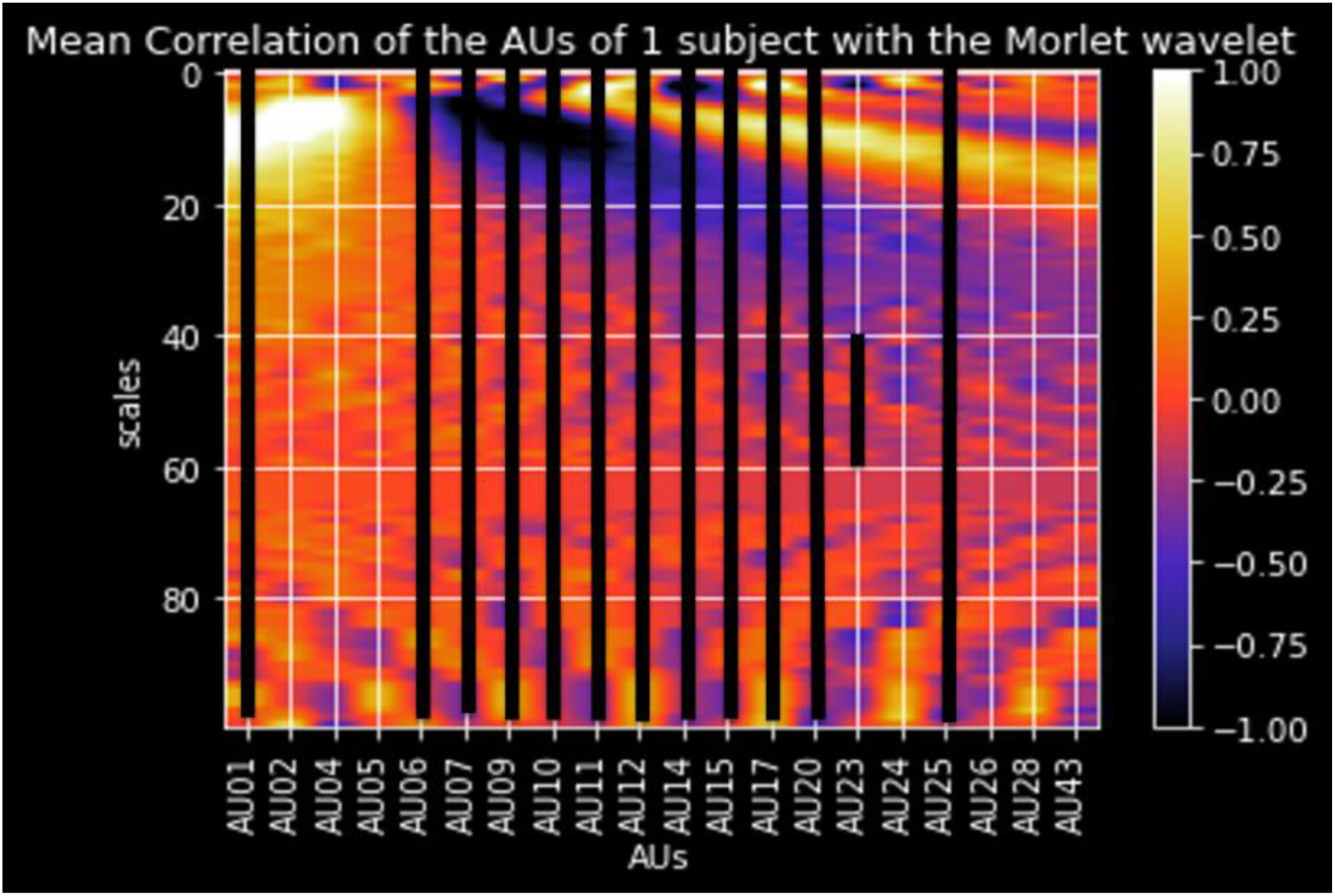

As for the advanced analysis we found 13 AUs in total to be statistically significant. Not all of them are mentioned in the literature currently but some of our results are in line with what previous studies found out, see in Discussion.

## 4. Discussion

Consistent with previous studies on the nonverbal expression of depression^22,23^, this study found that facial activities were associated with a depressive state. AUs can contribute crucial information about the mental state of an individual. As AUs capture information that cannot be given by questionnaires, they can be a better expression of the feelings instead of describing with words. Some AUs are more expressive than others.

This is an overview of the literature we found addressing specific action units:

| AU | description | Author | Quote/Findings |
| --- | --- | --- | --- |
| 1 | Inner Brow Raiser | Zhu et al. (2023)<sup>24</sup> | „our analysis showing decreased sadness with increased brow rotation, as FaceReader analysis correlates sadness with three facial action units: inner brow raiser, brow lowerer, and lip corner depressor”<br>(FaceReader = facial expression recognition software package)<br>[increased brow rotation = less expression of inner brow raiser → less expression of sadness], |
| 2 | Outer Brow Raiser |  |  |
| 4 | Brow Lowerer |  |  |
| 5 | Upper Lid Raiser | Blasberg et al. (2023)<sup>25</sup> | Occurrence associated with a higher level of psychosocial stress. |
| 6 | Cheek Raiser |  |  |
| 7 | Lid tightener | Blasberg et al. (2023)<sup>25</sup> | Occurrence associated with a higher level of psychosocial stress. |
| 9* | Nose Wrinkler | Kim et al. (2023)<sup>26</sup> | "participants with higher depressive symptom scores displayed more prominent facial muscle movements related to [...] wrinkled nose" |
|  |  | Liu et al. (2022)<sup>27</sup> | “we noted that AU9 (Nose Wrinkler) and AU10 (Upper Lip Raiser) were also closely related to CES-D scores, which are also representative of facial activities of disgust.” |
| 10 | Upper Lip Raiser | Liu et al. (2022)<sup>27</sup> | “we noted that AU9 (Nose Wrinkler) and AU10 (Upper Lip Raiser) were also closely related to CES-D scores, which are also representative of facial activities of disgust” |
|  |  | Blasberg et al. (2023)<sup>25</sup> | Occurrence associated with a higher level of psychosocial stress. |
| 11* | Nasolabial Deepener | Bernardes et al. (2021)<sup>19</sup> | The deepening of the nasolabial furrow is associated with pain in fetuses |
|  |  | Gilbert et al. (1999)<sup>20</sup> | Nasolabial Deepener is associated with postoperative pain in preschool children |
| 12 | Lip Corner Puller | Girard et al. (2014)<sup>28</sup> | “when symptom severity was high, AU 12 and AU 15 occurred less often. Participants smiled less and showed expressions related to sadness less often when severity was high.” |
|  |  | Blasberg et al. (2023)<sup>25</sup> | “more lip corner pulling (AU12) went along with lower cortisol reactivity” = less psychosocial stress |
|  |  | Girard et al. (2013)<sup>29</sup> | “Compared with low symptom severity interviews, high symptom severity interviews were marked by (1) significantly **lower overall AU 12 activity**, (2) significantly higher overall AU 14 activity” |
| 14 | Dimpler | Takemoto et al. (2023)<sup>30</sup> | „the intensity of Dimpler was higher in the control group than in the depression symptoms groups“ |
|  |  | Jung et al. (2023)<sup>31</sup> | “the HM component in MDD participants was inversely correlated with depressive symptoms on the BDI” (HM component = ”lip press“, ”**dimpler**“, ”lip suck“, and ”lip stretch“, representing lip-pursing/stretching movements”) |
|  |  | Girard et al. (2014)<sup>28</sup> | “AU 14 actually increased when participants were depressed” (so the opposite) |
|  |  | Girard et al. (2013)<sup>29</sup> | “Compared with low symptom severity interviews, high symptom severity interviews were marked by (1) significantly lower overall AU 12 activity, (2) significantly **higher overall AU 14 activity**” (also opposite to the two above) |
| 15 | Lip Corner Depressor | Girard et al. (2014)<sup>28</sup> | „AU 15, which is associated with both sadness and positive affect suppression, was decreased when depression was severe“<br>“when symptom severity was high, AU 12 and AU 15 occurred less often. Participants smiled less and showed expressions related to sadness less often when severity was high.” |
|  |  | Girard et al. (2013)<sup>29</sup> | „There were also two non-significant trends worth noting. Compared to the low symptom severity interviews, the high symptom severity interviews were marked by (4) **less overall AU 15 activity.**” |
| 17 | Chin Raiser | Takemoto et al. (2023)<sup>30</sup> | „Furthermore, in the Chin Raiser, the intensity is higher in the depression symptoms group when they were talking about negative conversation topics. It was supposed that participants in the depression symptoms group had negative emotional impacts from negative conversation topics more than in the control group, thus the intensity of Chin Raiser was higher.“ |
| 20* | Lip Stretcher | Jung et al. (2023)<sup>31</sup> | “the HM component in MDD participants was inversely correlated with depressive symptoms on the BDI” (HM component = ”lip press“, ”dimpler“, ”lip suck“, and ”**lip stretch**“, representing lip-pursing/stretching movements”) |
| 23 | Lip Tightener |  |  |
| 24* | Lip Pressor | Kim et al. (2023)<sup>26</sup> | “increased expression in facial muscles involved in blinking, depressed, and tight lip movements in participants with higher depressive symptom scores” |
|  |  | Jung et al. (2023)<sup>31</sup> | “the HM component in MDD participants was inversely correlated with depressive symptoms on the BDI” (HM component = ”**lip press**“, ”dimpler“, ”lip suck“, and ”lip stretch“, representing lip-pursing/stretching movements”) |
| 25* | Lips Part |  |  |
| 26* | Jaw Drop |  |  |
| 28 | Lip Suck | Jung et al.<br>(2023)<sup>31</sup> | “the HM component in MDD participants was inversely correlated with depressive symptoms on the BDI” (HM component = ”lip press“, ”dimpler“, ”**lip suck**“, and ”lip stretch“, representing lip-pursing/stretching movements”) |
| 43* | Eyes Closed | Hinchliffe<br>et al.<br>(1971)<sup>32</sup> | Depressed patients make less eye contact than controls or recovered patients |

In 2023, Kim et al. found that “participants with higher depressive symptom scores displayed more prominent facial muscle movements related to […] wrinkled nose”^26^, which is in line with what we found out about AU9. In general, this AU is associated with disgust. This aligns with the observation that feelings of depression and self-disgust frequently appear together and consistently, affecting 80% or more of patients with depression^33^. The study by Kim et al. included a dataset of patients with an average age of 71.98 years, making it interesting to see similar results in younger patients. Liu et al. noted that the expression of AU9 was closely related to a higher CES-D (Center for Epidemiological Studies Depression) score, which is another commonly used depression questionnaire^27^. These sources underline the importance of this specific AU.

Our analysis of AU11 revealed a negative correlation with the BDI score, indicating that higher depression scores were associated with a reduced expression of pain in the subjects’ faces. Interestingly, previous studies on children and fetuses have linked the deepening of the nasolabial furrow (AU11) to pain expression^19,20^, suggesting that in depressed individuals, the facial signals of pain may be reduced despite the association of this action unit with pain in other populations.

For AU 24 we found a positive correlation with the BDI score, meaning that patients with more severe depressive symptoms happen to have their mouth closed more often, or more specifically, press their lips against each other. Kim et al. also suggested an “increased expression in facial muscles involved in […] depressed and tight lip movements in participants with higher depressive symptom scores” ^26^.

FACS associates AU25 mainly with the emotion happiness, and also surprise and fear. It is expected to be expressed less the higher the BDI score is, which is in line with our findings. According to FACS happiness is associated with the AUs 1, 12 and 25, while 12 (Lip Corner Puller) and 25 (Lips Part) are the two that produce the motion of an open smile.

According to FACS, the muscles associated with AU26 contribute to the emotions fear and surprise. This AU seems to be expressed less the higher the BDI score. As AUs 25 and 26 both play a significant role in the expression of fear and surprise, these two emotions seem to be expressed less by patients with depressive symptoms.

AU 43 is a behavioral AU and we found it to be positively associated with the BDI score. Studies have shown that individuals with depression tend to open their eyes less widely, blink for longer durations^34^, and make less eye contact^32^ compared to those in the general population or those who have recovered. All of these states go along with the motion of closing the eyes more in patients with high depressive scores, and it is in-line with our results.

Digital information sources provide access to valuable, naturally occurring health-related medical information. This can enable real-time monitoring of symptoms to support diagnosis and treatment recommendations. Compared to existing precision psychiatry, the practice uses static information.

Recent advancement in digital health technologies is the development of the digital twin, which is a simulation of a system (i.e., patient’s dynamic mental state) that uses the best available data/features/models to mirror the system and predict its future states. This can be achieved by using real world data combined with advanced mathematical tools.

Then, the prediction outcome will be for the actual person and not for the average person resulting to the absolute precision psychiatry. Such technologies will address the problem with scarce data in psychiatry as the prediction model will be for the exact person.

The aim is to develop advanced digital tools that enable intensive dynamic tracking of symptoms and behaviour and use them to predict short-and long-term outcomes. Thus, clinicians will spot patterns that point to illness deterioration or improvement even in real time.

Looking forward, it is necessary to validate the findings by increasing the sample size of the dataset.

Some limitations must be acknowledged in this research. Variations in the recording environment, such as differences in lighting, background, and camera angles, can compromise the quality of video data, making it challenging to detect consistent patterns. Additionally, patients may behave differently in a clinical setting compared to their natural environment, which can result in unnatural facial expressions and body movements. Psychiatric symptoms can vary widely across different cultures and individual backgrounds, making it essential to have a larger and more varied dataset to accurately capture this diversity and improve the model’s applicability.

## 5. Conclusion

This study is one of the first to look at how facial expressions relate to depressive symptoms across different psychiatric diagnoses. The results show that facial expressions from short video recordings are closely linked to the severity of depression. By combining facial expression analysis with advanced mathematical tools, this approach could help detect depression earlier and create more personalized treatment plans. This may lead to better outcomes for patients through faster and more personalized care. Overall, this research is an important first step in studying how facial expressions connect with depressive symptoms transdiagnostically.

## Data Availability

All data produced in the present study are available upon reasonable request to the authors

## Acknowledgments

The equipment used for the recordings was funded by the UKSH Research Foundation in 2022 (https://www.uksh.de/gutestun/foerderstiftung.html).

Prof. Giorgos Giannakakis and Prof. Anastasios Roussos from the FORTH institute in Greece supported on the set up of the lab used for the clinical interviews.

## Notes

### Competing Interest Statement

The authors have declared no competing interest.

### Author Declarations

Ethics committee of the University of Luebeck gave ethical approval for this work on the 19th July 2024 (file number 2024-355_1)

